# Enhancing patient representation learning from electronic health records through predicted family relations

**DOI:** 10.1101/2024.03.12.24304163

**Authors:** Xiayuan Huang, Jatin Arora, Abdullah Mesut Erzurumluoglu, Daniel Lam, Boehringer Ingelheim – Global Computational Biology and Digital Sciences, Hongyu Zhao, Zhihao Ding, Zuoheng Wang, Johann de Jong

## Abstract

Artificial intelligence and machine learning are powerful tools in analyzing electronic health records (EHRs) for healthcare research. Despite the recognized importance of family health history, in healthcare research individual patients are often treated as independent samples, overlooking family relations. To address this gap, we present ALIGATEHR, which models predicted family relations in a graph attention network and integrates this information with a medical ontology representation. Taking disease risk prediction as a use case, we demonstrate that explicitly modeling family relations significantly improves predictions across the disease spectrum. We then show how ALIGATEHR’s attention mechanism, which links patients’ disease risk to their relatives’ clinical profiles, successfully captures genetic aspects of diseases using only EHR diagnosis data. Finally, we use ALIGATHER to successfully distinguish the two main inflammatory bowel disease subtypes (Crohn’s disease and ulcerative colitis), illustrating its great potential for improving patient representation learning for predictive and descriptive modeling of EHRs.

## Introduction

Recent years have seen a surge of interest in utilizing electronic health records (EHRs) for healthcare research^1,2^, including modeling disease risk, onset, and progression^3-6^. EHR databases store data routinely collected in a primary and/or secondary care setting and are a comprehensive resource of a patient’s disease history, by providing historical information on diagnoses, prescriptions, laboratory tests, (surgical) procedures, and doctor’s notes.

Understanding an individual’s family health history is critical in healthcare and medicine, for example, assessing patients’ risk of common and rare diseases such as heart disease, type 2 diabetes, and cancer. This is because families share genetic variations, environmental exposures, and social determinants of health^7,8^. However, EHR research so far has adopted a limited view of family relations at best, essentially treating individual patients as independent samples. One reason for this is that family histories are not systematically recorded in EHR databases. Family-related information is largely captured via survey or active questioning by healthcare professionals^9^ and stored in EHRs as free text or scanned documents, limiting its usability in healthcare research^10^. For this reason, several rule-based algorithms have recently been developed for inferring electronic family pedigrees (e-pedigrees) from EHR data^11-13^. These methods use basic demographic and/or emergency contact information readily available in most EHRs, and their application across a wide spectrum of diseases have provided strong evidence for the general usability of e-pedigrees in genetic and epidemiological research. Meanwhile, their results show that a dataset with more than one million individuals can be adequately used to infer entire families^12,14^.

Due to their ability to extract patterns from large and complex datasets such as EHR databases, artificial intelligence and machine learning have shown great promise in modeling EHR data as well^15,16^. In one of the earliest attempts, “Doctor AI” used recurrent neural networks to predict medical events such as diagnoses, medications, and procedures, from historical EHR data^17^. Since then, deep learning methods such as word embedding^18^, graph machine learning^19-21^, graph-based attention models^19,22^, and patient representation learning^23-25^ have demonstrated great potential for a variety of healthcare-related tasks. Despite the significant advancements in the analysis of EHRs using machine learning, all existing methods still treat individual patients as independent samples without considering family relations. Explicitly incorporating family health history could benefit a wide range of tasks, including modeling disease risk, onset, and progression, as well as patient segmentation. For example, much like a physician leveraging knowledge of family health history to assess a patient’s risk of cardiovascular disease, an explicit representation of family health history could enhance the predictive accuracy of a machine learning model in forecasting such risks.

In this work, we address this critical limitation in the current utilization of EHRs for healthcare research by integrating the recent progress in family pedigree prediction with the parallel progress in using machine learning for analyzing EHR data. We present ALIGATEHR (ALIgning Graph Attention neTworks for EHR), a generic framework for learning patient representations, which models predicted family pedigrees using a graph attention network of long short-term memory (LSTM) nodes^26^. To further enhance the quality of the learned representations, we additionally integrate a medical ontology of diagnosis codes into the attention mechanism. Taking disease risk prediction as a use case, we demonstrate that explicitly modeling family relations markedly improves performance across thousands of diagnosis codes, as compared to state-of-the-art baseline methods. We then demonstrate the interpretability of our model using an attention-based feature importance score^27^, allowing us to quantitatively assess the impact of the health history of family members on the disease risk of a patient. The pedigree-based attention mechanism enables ALIGATEHR to capture genetic aspects of diseases using only EHR data as input. Finally, we show that the patient representations learned by ALIGATEHR can successfully distinguish between two major subtypes of inflammatory bowel diseases (IBD), Crohn’s disease (CD) and ulcerative colitis (UC), using only EHR data as input. Our results demonstrate that the importance of explicitly incorporating family relations in EHR modeling should not be overlooked, and that ALIGATEHR can serve as a powerful tool for improving the quality of patient representations in a variety of downstream predictive and descriptive tasks.

## Results

### Overview of ALIGATEHR

Our proposed model, ALIGATEHR, aims to explicitly capture dependencies between related patients and diseases from EHRs and medical ontologies, to learn more informative patient representations that can be utilized for a variety of downstream tasks. ALIGATEHR, by design, focuses on the sequential order of visits for each patient, without considering the temporal intervals between visits. In ALIGATEHR, a patient’s EHR trajectory is modeled using a recurrent neural network. Related patients are identified via the E-Pedigrees software^11^ and connected via an attention mechanism^28^ into a patient-level graph attention network (GAT). An attention mechanism is also used to connect the patient-level GAT to a medical ontology, to allow for capturing dependencies between diseases. Details of the architecture of ALIGATHER are provided in Fig. 1 and the Methods section.

**Fig. 1:**
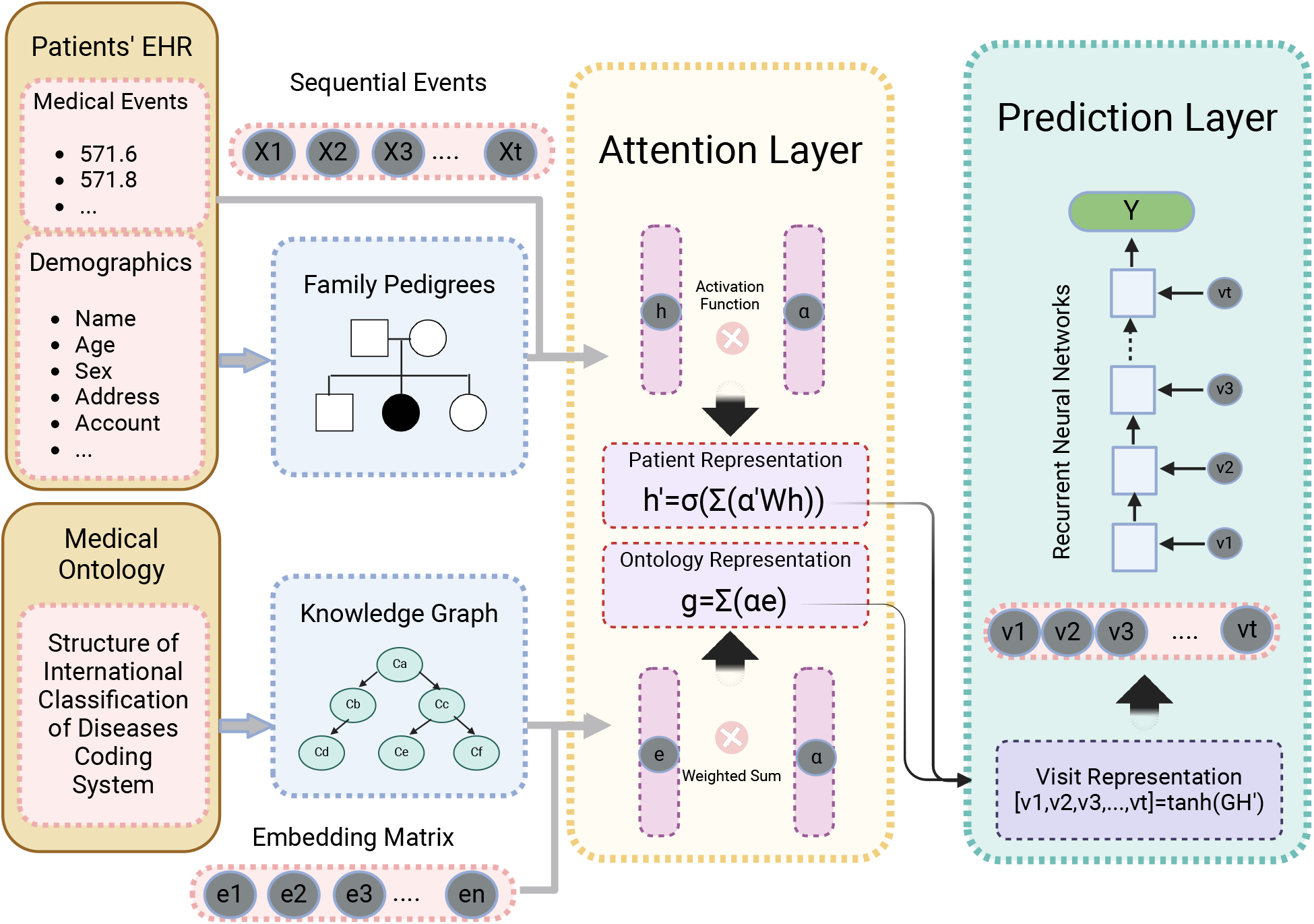
Illustrative diagram of ALIGATEHR. ALIGATEHR consists of two parallel processes: 1) Construction of pedigree graph: the patient representation *h*′ aggregates health information from relatives; and 2) Construction of ontology graph: the representation *g* captures dependencies between diseases. The final representation *ν*_*t*_ merges both patient and ontology information to represent a patient’s disease status for each visit. Finally, a series of visits is fed into a neural network model for the risk prediction task.

### ALIGATEHR outperforms baseline models on risk prediction across the disease spectrum

We evaluated the performance of ALIGATEHR in a risk prediction setting, using data from the Merative™ MarketScan® Research Databases, which include longitudinal medical records of over 660,000 individuals involved in predicted families with an average family size of 2.4 (Extended Data Fig. 1 and Extended Data Table 1). Our prior studies have validated that E-Pedigrees construct family pedigrees with high probability, resulting in a very low false positive rate^11,14^. This is primarily attributed to the conservative nature of the E-Pedigrees algorithm, ensuring the construction of family relations with high confidence. This conservative approach enhances reliability and precision for downstream analysis. The predicted families have demonstrated substantial utility in disease risk prediction and genetic research. To construct pedigree networks, we captured first-degree relatives for each person in the pedigree. In our analysis, we included all patients with at least 2 visits, and the average number of visits per patient was 7.9. The dataset contained 13,097 unique ICD-9 codes, but a significant portion of these were infrequently used for clinical diagnosis, either due to redundancy or the low prevalence of the corresponding diseases. This dataset also contained ICD-9 codes unrelated to diseases, as well as those beginning with the letters ‘E’ or ‘V’. E-codes detail the external factors leading to diseases or injuries, whereas V-codes serve purposes unrelated to diseases or injuries, instead providing supplementary documentation information. We excluded ICD-9 codes of low prevalence, E-codes, V-codes, and ICD-9 procedure codes from consideration. After data processing, we treated risk prediction as a binary classification problem to predict whether a given patient would receive a specific diagnosis during the next clinical visit. We constructed a total of 1,886 predictive models, each corresponding to a specific diagnosis code. It is worth noting that a family member was considered positive for a diagnosis code only if that code appeared two or more times in that family member’s health record, following the “rule of two”^29^.

We then compared ALIGATEHR with state-of-the-art models, including:

1. Classical machine learning models: Logistic Regression (LR) and eXtreme Gradient Boosting (XGBoost)^30^,
2. Recurrent neural networks: Long Short-Term Memory (LSTM),
3. Skip-gram based Models^31^: Med2Vec^18^,
4. Attention-based models: GRAM^22^ and Dipole^32^.

Averaged across all diseases, ALIGATEHR showed superior performance compared to all baseline models (Table 1a). The average area under the curve (AUC) of ALIGATEHR over all 1,886 diseases was 0.843, significantly higher than the second-largest average AUC of 0.809 among all methods applied (Wilcoxon signed-rank test, P=1.95E-303). ALIGATEHR outperformed classical machine learning by more than 30*t* and other neural network-based models by at least 5*t*. Moreover, ALIGATEHR outperformed all other methods for almost all individual diseases (Fig. 2a). The most substantial improvements were observed for diseases in the ICD diagnosis groups of “Neoplasms”, “Endocrine, nutritional and metabolic diseases”, “Diseases of the genitourinary system” and “Diseases of the respiratory system” (Fig. 2b), suggesting that family health history and relations play an important role in conferring disease risk in these four diagnosis groups. As an example, many diagnoses in the “Neoplasms” group are known to exhibit significant familial clustering^33^, which explains the substantial improvement observed when integrating family health history into risk prediction modeling. Specifically, within the “Neoplasms” group, the five most significantly improved diseases compared to the baselines are “Nodular lymphoma”, “Malignant neoplasm of brain”, “Malignant neoplasm of thyroid gland”, “Malignant neoplasm without specification of site”, and “Multiple myeloma”. This observation can be attributed to the heightened risk of developing these diseases when there is a first-degree relative with a similar medical history^34-37^. Interestingly, the smallest improvement was observed in the diagnosis group of “Injury and Poisoning”. Although overall, ALIGATEHR still significantly outperformed all other methods in this group, there are individual diagnoses where the difference was less clear: Dipole achieved marginally better performance than ALIGATEHR for 26 individual codes (such as “Injury of chest wall”, “Fracture of ankle” and “Contusion of thigh”) out of the 273 codes, and GRAM for 5 individual codes (such as “Acute myocardial infarction of anterolateral wall” and “Lung contusion”) out of 273 codes. Likely, ALIGATEHR loses some of its competitive advantage in settings where family relations (and the genetic heritability that these capture) are less important, such as accidents, environmental hazards, and lifestyle choices, all typical diagnoses in the “Injury and Poisoning” group. These results highlight the substantial impact that modeling family health history and relations can have on tasks such as disease risk prediction.

**Table 1:**
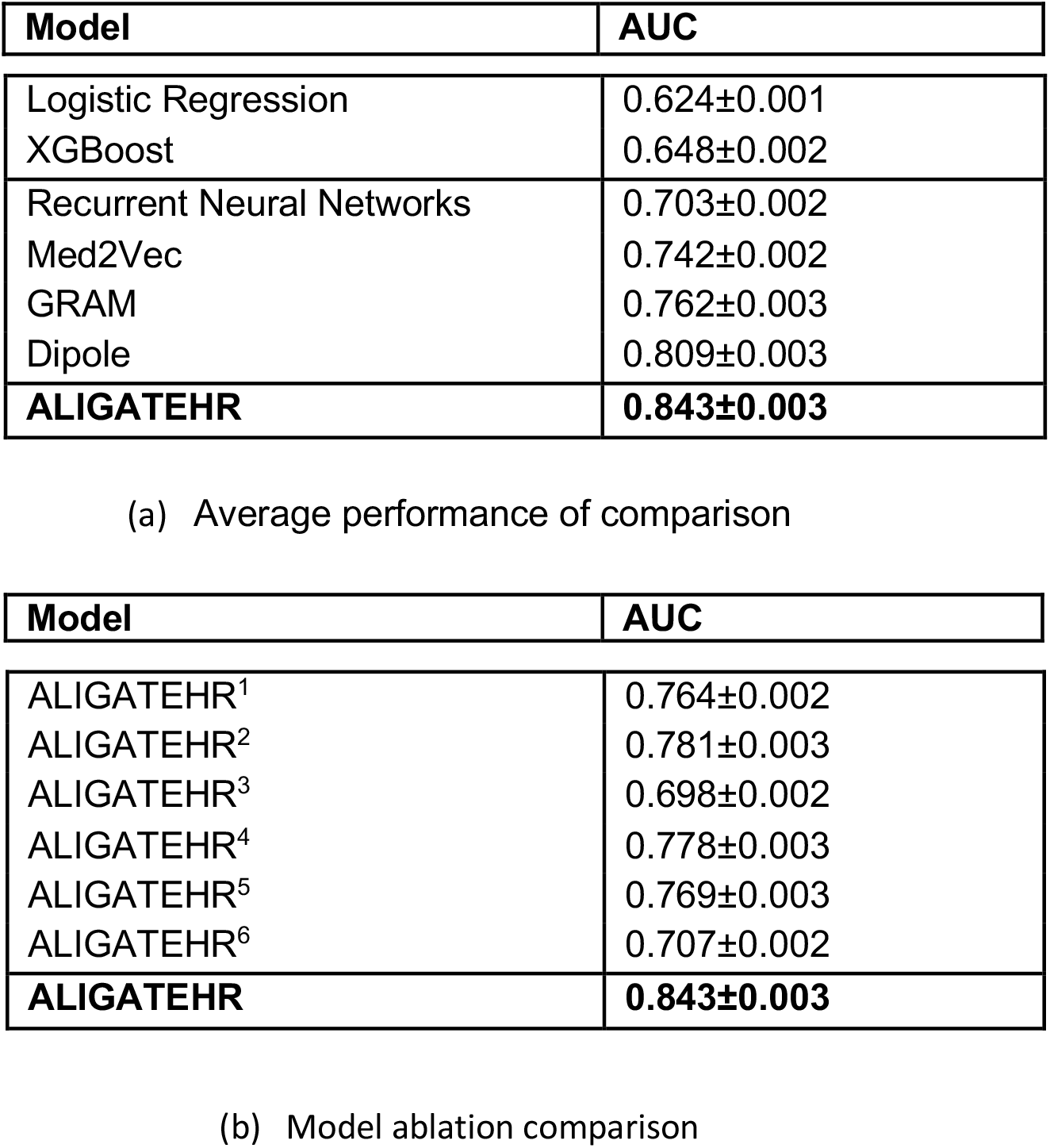
Comparison of model performance. (a) Average performance on disease risk prediction tasks with 95% confidence interval. (b) Model ablation comparison: ALIGATEHR^1^ without pedigree graph; ALIGATEHR^2^ without ontology graph; ALIGATEHR^3^ without both pedigree graph and ontology graph; ALIGATEHR^4^ with constant weights on edges of the pedigree graph; ALIGATEHR^5^ with constant weights on edges of the ontology graph; ALIGATEHR^6^ with constant weights on edges of both pedigree graph and ontology graph.

**Fig. 2:**
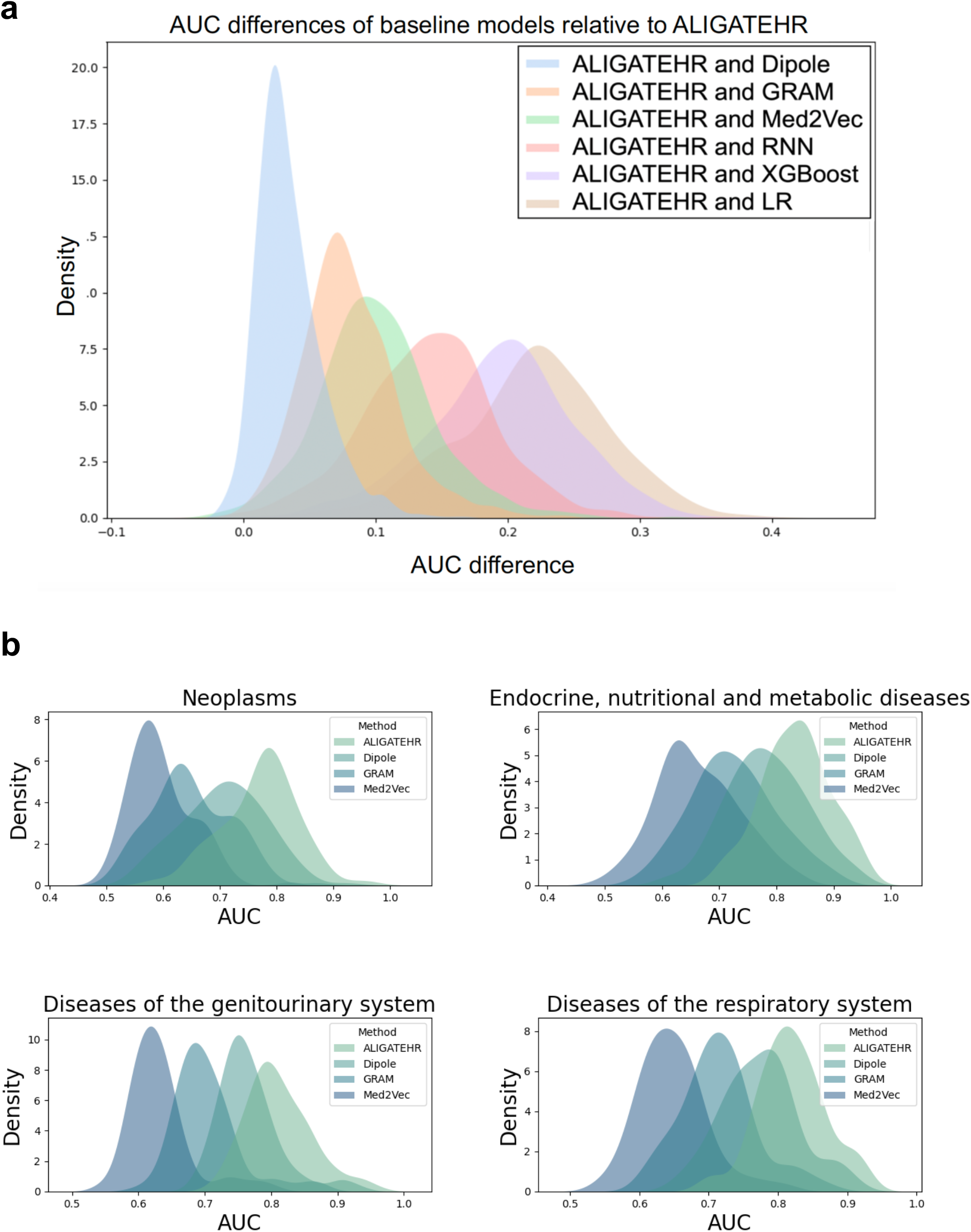
Performance evaluation of ALIGATEHR using area under the curve (AUC). **a**, Kernel density estimate (KDE) of the AUC difference between ALIGATEHR and six baseline models across all diseases, showing that ALIGATEHR outperforms all other methods for almost all individual diseases. **b**, KDE of AUC distribution for ALIGATEHR, Dipole, GRAM, and Med2Vec, in four ICD diagnosis groups where ALIGATEHR has the most improvement.

### ALIGATEHR relies on all model components for achieving its superior performance

To gain insights into how its individual components contribute to the performance of ALIGATEHR, we performed an ablation study, sequentially removing or disabling its individual components. Most importantly, we observed that the removal of any individual component significantly affected the performance of ALIGATEHR (Table 1b). Of all individual ablations, removing the e-pedigree attention mechanism from ALIGATEHR degraded performance the most, again highlighting the importance of incorporating family relations in risk prediction models. As expected, this led to a performance (AUC = 0.764) that was very close to that of GRAM (AUC = 0.762), given that ALIGATEHR without e-pedigrees is architecturally highly similar to GRAM. Interestingly, removing the e-pedigree attention mechanism altogether led to worse performance than using constant e-pedigree attention weights (in essence a graph convolutional network). This means that just connecting patients with their relatives in a graph is insufficient for achieving optimal performance (constant e-pedigree attention weights). Optimal performance is only achieved when we enable selecting the right family members from among all relatives (trained e-pedigree attention weights).

### ALIGATEHR is highly interpretable and captures genetic aspects of diseases

Having assessed the performance of ALIGATEHR in a risk prediction setting, we then wanted to assess its interpretability using an attention-based feature importance score. Attention-based feature importance allows us to quantify the impact of family members’ diagnosis histories on a patient’s risk prediction outcome. For this purpose, we employed GNNExplainer^27^, which was designed to provide interpretable explanations for predictions in graph neural networks and has robust performance in examining attention-based graph neural networks through producing scores that highlight the significance of features. Fig. 3a shows the ranking of features, based on their feature importance scores averaged across all diseases, thus representing the importance of features to general human health. As expected, we found many common diseases associated with general human health among the top 200 features, such as “Pure hypercholesterolemia”, “Obesity”, “Hyperlipidemia”, “Essential hypertension”, “Hyperosmolality and/or hypernatremia” and “Anemia”. Interestingly, we also found several mental health disorders, including “Anxiety”, Depressive disorder”, “Dysthymic disorder” and “Bipolar I disorder”, were ranked among the top 200 features. This indicates that having a family history, particularly with first-degree relatives affected by mental health disorders, is associated with an increased risk for an individual’s general health.

**Fig. 3:**
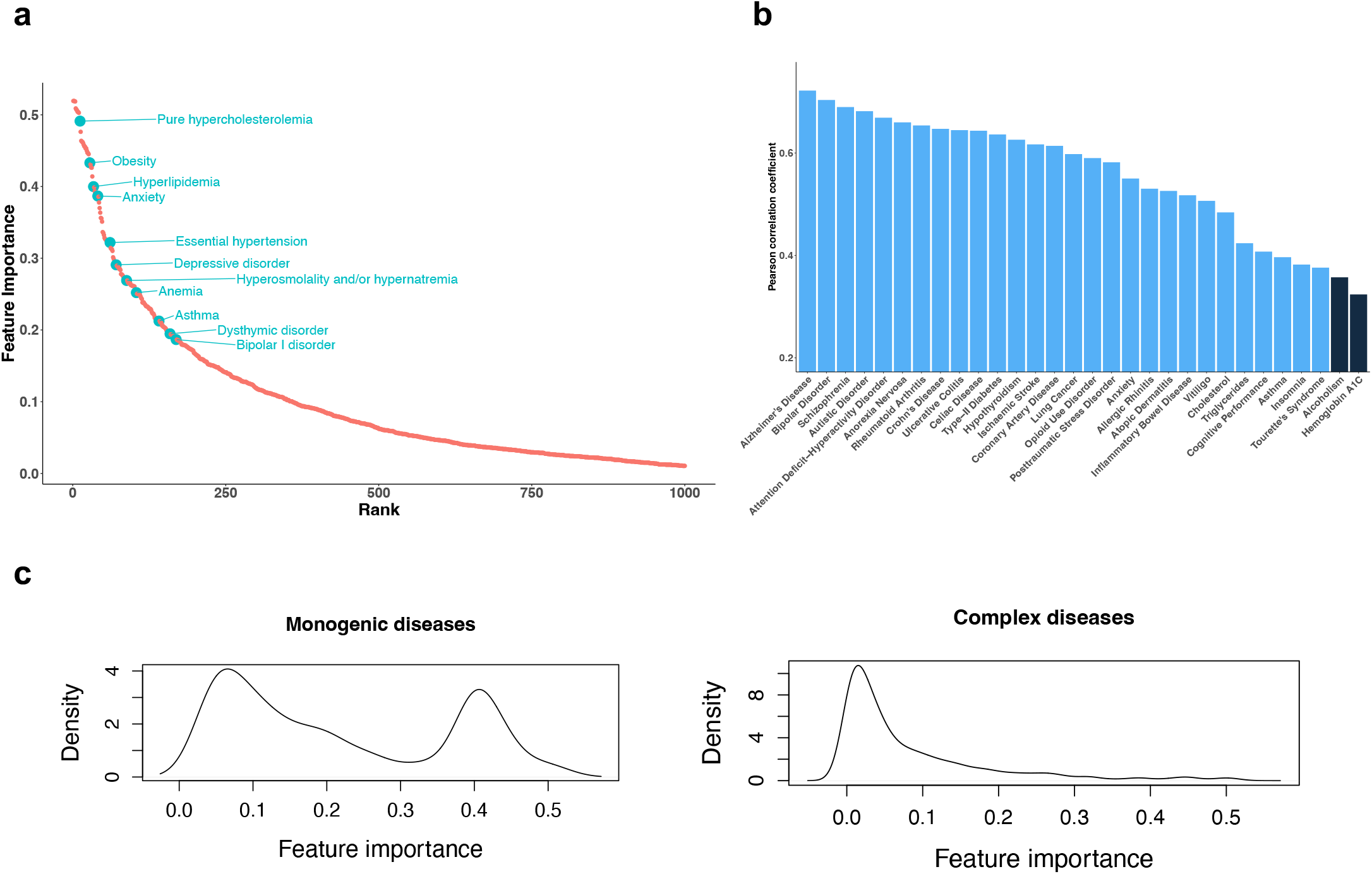
Interpretability analysis of ALIGATEHR in capturing genetic aspects of diseases. **a**, Feature importance ranking based on the feature importance scores averaged across all diseases, representing the importance of features to general human health. The x-axis represents the ranking of each feature; the y-axis represents the averaged feature importance. Highlighted diseases are common conditions or risk factors associated with general human health. **b**, Pearson correlation of genetic correlation (between features and the outcome) and feature importance (associated with the outcome) for 30 disease outcomes. Diseases in blue bars showing significant correlation with a false discovery rate (FDR) < 0.05. **c**, Kernel density estimate (KDE) of feature importance for monogenic and complex disease groups, where bimodality in the monogenic disease feature importance score distribution and right-skewed in the complex disease feature importance score distribution.

To investigate to what extent ALIGATEHR’s attention mechanism captured information relevant to disease biology, we curated a set of 30 prevalent diseases with known genetic correlations with each other, sourced from LD Hub^38^, following the exclusion of traits without corresponding ICD codes or those not present in EHRs such as “Height” and “Educational attainment”. For each disease outcome, we retrieved the importance scores of features associated with the outcome, as well as the genetic correlations between features and the outcome. We then computed the Pearson correlation coefficient between the genetic correlation and the feature importance (Fig. 3b). Among the 30 diseases we considered, 28 disease outcomes had a significant correlation with a false discovery rate (FDR) < 0.05. This means that features exhibiting a strong genetic correlation with a certain disease outcome play a significant role in influencing the risk of that disease in our proposed model and demonstrates that ALIGATEHR is highly effective at capturing genetically relevant information using its pedigree-based attention mechanism. Interestingly, we again found many mental health-related diseases among the most strongly correlated diseases, including “Bipolar disorder (BPD)”, “Schizophrenia (SCZ)”, “Autistic disorder (AUT)”, and “Attention deficit-hyperactivity disorder (ADHD)”. These diseases were previously reported with high heritability (BPD: 60-80*t*; SCZ: 70-80*t*; AUT: 70-90*t*; ADHD: 70-80*t*)^39-42^.

To further investigate the extent to which ALIGATEHR’s attention mechanisms captured genetic aspects of disease, we turned to the distinction between monogenic diseases and complex diseases. A monogenic disease results from mutations in a single gene, whereas complex diseases involve the cumulative impact of genetic variants in multiple genes and their interactions with the environment. We compiled a set of 14 monogenic diseases such as “Cystic fibrosis”, “Familial hypercholesterolemia”, and “Muscular dystrophy”, from the literature. We trained risk models for all monogenic diseases and averaged the resulting feature importance scores to arrive at a single list of feature importance scores for the group of monogenic diseases. We treated the remaining 1,872 diseases as complex diseases, and similar to above, computed a single list of feature importance scores representing the group of complex diseases. Remarkably, we observed that for the monogenic diseases, the major risk factors of disease could be clearly identified by the magnitude of their feature importance score, as illustrated by the bimodality in the kernel density estimate of the monogenic disease feature importance score distribution (Fig. 3c). On the other hand, complex diseases seemed to be more diffusely influenced by a multitude of risk factors, as indicated by the right-skewed distribution with a peak around 0.01. These results verified that ALIGATEHR, despite using only EHR data, can capture aspects of the genetic architecture of diseases.

### Application: distinguishing inflammatory bowel disease subtypes

To illustrate the utility of ALIGATEHR in analyzing individual diseases, we applied ALIGATEHR to the problem of distinguishing the subtypes of inflammatory bowel disease (IBD) based on EHR data only. IBD is a chronic disorder that involves inflammation of the gastrointestinal (GI) tract. Its burden is increasing worldwide^43^ due to the increasing prevalence and incidence rate over the past few decades^44^ and the tendency of IBD to frequently occur with comorbidities^45^. This public health concern is projected to escalate in the coming years, driven by the strongest risk factor of having a relative with the disease^46^. Crohn’s disease (CD) and ulcerative colitis (UC) are the two major subtypes of IBD and are primarily distinguished by location within the GI tract where the inflammation occurs^47^. Genetic studies have identified many susceptibility loci for IBD, mostly shared between CD and UC^48^. The genetic correlation between CD and UC is 0.634^38^. In clinical practice, differentiating between CD and UC can be challenging due to the overlap in symptoms, leading to misdiagnosis rates of up to 10*t*^49^. This similarity in clinical presentation was reflected in our analyses. First, CD and UC demonstrated comparable Pearson correlation coefficients of 0.647 (P=1.48E-4) and 0.645 (P=1.60E-4), respectively (Fig. 3b), which suggested a shared set of risk factors and genetic components between these IBD subtypes. This was further supported by the observation that 16 features were shared among the top 20 features for CD and UC (Fig. 4a). Hence, we wondered if we could use ALIGATEHR to distinguish CD and UC as distinct IBD subtypes from EHR data only. For this purpose, we merged the CD and UC patient populations (n = 2,324 and n = 1,651, respectively) and trained our model for predicting the risk of overall IBD. It appears that in the analysis of patient representations obtained from the model, ALIGATEHR successfully distinguishes between CD and UC patients (Fig. 4b). A small number of patients were mixed or overlap near the boundary of the two clusters. This could reflect the challenges in differentiating between CD and UC in clinical practice, as mentioned above. Interestingly, 2.8% of UC patients (n = 47) appeared to be distinctly clustered within the CD group, i.e. had a clinical presentation that is very much like CD. These patients could represent cases of CD misdiagnosed as UC in clinical practice. In conclusion, despite the shared risk factors and symptoms between CD and UC, ALIGATEHR is highly effective at distinguishing between the two conditions using only EHR data as input.

**Fig. 4:**
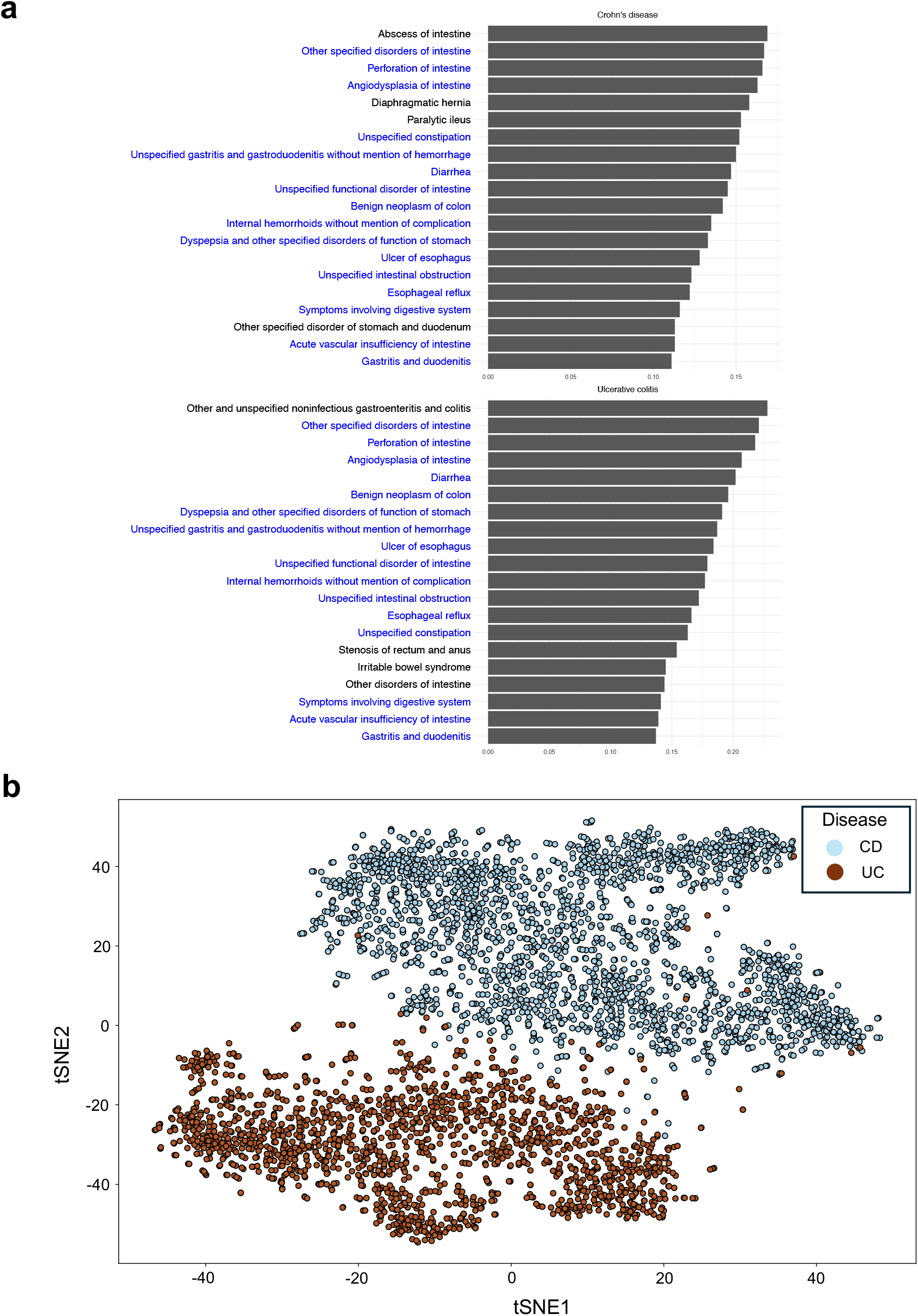
Interpretability analysis of ALIGATEHR in distinguishing inflammatory bowel disease (IBD) subtypes. **a**, Top 20 feature importance ranking for Crohn’s disease (CD) and ulcerative colitis (UC), showing 16 features are shared between CD and UC. The x-axis represents feature importance; the y-axis represents conditions, where conditions in blue are common features for both CD and UC. **b**, t-SNE plot of IBD patients using learned latent patient representation, showing the separation and clustering of CD and UC subtypes.

## Discussion

EHR databases store health-related information from millions of patients, routinely collected over many years in primary and/or secondary care settings. EHR data provides a comprehensive history of patients and their diseases by covering multiple modalities, such as diagnoses, prescriptions, and lab tests. In recent years, significant advancements have been made in developing artificial intelligence and machine learning methods to effectively manage the size and complexity of EHR data for improving our understanding of patients and diseases. Despite this progress, the well-documented influence of family relations has been largely overlooked.

In this work, we addressed this important limitation by presenting ALIGATEHR, a deep learning framework for integrating inferred family pedigrees as edges into a GAT, where the nodes are patient-specific EHR trajectories modeled using LSTM networks. Additionally, a medical ontology is integrated into ALIGATEHR via an attention mechanism. We showed that explicitly modeling family relations using ALIGATEHR leads to substantial performance gains in a risk prediction case study. Through ablation experiments, we demonstrated that all components of ALIGATEHR are essential for achieving its optimal performance. Moreover, we showed that ALIGATEHR is highly interpretable, and can capture genetic aspects of diseases using its attention-based feature importance scoring, without using any genetic data as input. Finally, we showed that the patient representations learned by ALIGATEHR can successfully distinguish between two closely related IBD subtypes, CD and UC, using only EHR data as input.

A limitation of this study lies in the incomplete nature of EHRs, where only a fraction of patients could be linked to family pedigrees. Another potential limitation arises from the structure of the predicted families. The E-Pedigrees software employs a conservative rule-based decision tree algorithm for family relation inference, often resulting in the construction of a substantial number of small-sized families with a limited number of siblings within each family. Despite these incomplete e-pedigree data and small families, our experiments still yielded significant improvements in performance, underlining the importance of including family health history in predictive healthcare models. Future work that can acquire more comprehensive family health history may further improve the predictive power.

In this work, our primary focus was on disease risk prediction. We achieved this by incorporating health history from relatives before the disease manifestation in the patient, through attention-based pedigree graphs and recurrent neural networks. It provides a more comprehensive understanding of how a patient’s previous health records and family health history collectively impact the disease risk for that individual. It is important to note that ALIGATEHR is a generic framework for patient representation learning. Applications of ALIGATEHR are not restricted to disease risk prediction but include a wide range of downstream predictive and descriptive tasks, such as modeling disease onset and progression, as well as patient segmentation. For example, in patient segmentation, the attention-based feature importance scores could help in identifying novel patient subgroups for a given disease, based specifically on patterns of family-related risk factors. This could open up new opportunities for developing more personalized treatment plans and prevention strategies, enhancing the effectiveness of healthcare interventions.

In addition to expanding the number of downstream applications beyond disease risk prediction, future research could explore the redefinition of edges in ALIGATEHR. While current edges are defined based on family relations, alternative measures of patient similarity may be more appropriate depending on the specific use case. For example, in biobanks such as the UK Biobank^50^, it is not always feasible to infer family relations between individuals, either because family information is unavailable or because the biobank cohort is too sparsely sampled from the general population. However, biobanks often provide additional data modalities, allowing the possibility of defining edges between patients based on measures of (disease-specific) genetic similarity. The inclusion of such additional data types would likely enhance the model’s ability to interpret and quantify the interplay between various risk factors, and further improve our understanding of diseases.

In summary, our results show that ALIGATEHR has the potential to substantially enhance patient representation learning for a variety of downstream predictive and descriptive tasks.

## Methods

### Notations

We denote the set of entire diagnosis codes from EHR as *d*_1_, *d*_2_, …, *d*_|*C*|_ ∈ *C* with the vocabulary size |*C*| (detailed description of notations is in Extended Data Table 2). The health record of each patient can be described as a sequence of visits *V*_1_, …, *V*_*t*_, where each visit contains a subset of diagnosis codes *V*_*t*_ ⊆ *C. V*_*t*_ can be represented as a binary vector *X*_*t*_ ∈ {0,1}^|*C*|^ in which the *k*-th element is 1 if *V*_*t*_ contains the code *d*_*k*_. In the context of a given medical ontology 𝒢, the hierarchy of various medical concepts is often represented through a parent-child relationship, with the diagnosis codes *D* serving as the leaf nodes, *D* = {*d*_1_, *d*_2_, …, *d*_|*D*|_} where |*D*| is the number of all leaf nodes. Ontology *𝒢* is depicted as a directed acyclic graph (DAG) in which the nodes constitute a set *C* = *D* + *D*′. The set 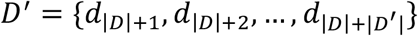 represents the collection of non-leaf nodes, where |*D*′| denotes the number of all non-leaf nodes. We refer to the DAG representation of 𝒢 as a knowledge graph of medical ontology. In the knowledge DAG, a parent node represents a more general medical concept than its children. Consequently, *𝒢* offers a multi-level perspective on medical concepts, presenting varying levels of specificity. This allows for a comprehensive understanding of the medical domain, accommodating concepts at different levels of granularity.

### Attention mechanism on pedigrees

Family pedigrees are inferred via the E-Pedigrees software^11^ using available demographic information from EHR. A graph of patients is then built by connecting via the inferred first-degree relations in the family. Each node (patient) in the graph has an associated feature vector representing a patient’s disease status, denoted as *h*_*i*_ ∈ ℝ^*C*^, with |*C*| the total number of unique diagnosis codes in the EHR data. Between the nodes, an attention mechanism is used to link a patient’s representation to the clinical profiles of his/her relatives. We formulate the patient’s final disease representation as follows:

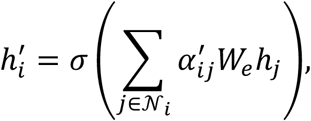

where 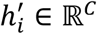 denotes the final representation of patient *i, σ* applies a ReLU activation function, 𝒩_*i*_ contains the indices of patient *i*’s first-degree relatives, *W*_*e*_ is the weight matrix which is applied to each node for a shared linear transformation, and the attention coefficient 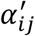 of e-pedigrees graph is calculated by the following softmax function,

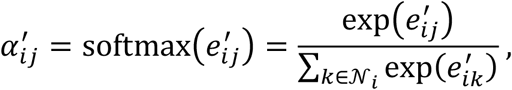

where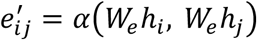. In our experiments, the attention mechanism is a single-layer feedforward neural network. Here 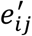 indicates the importance of first-degree relative *j*’s disease features to patient *i*.

### Attention mechanism on knowledge graph

In the ontology DAG, each node is assigned a basic embedding vector *e*_*i*_ ∈ ℝ^*m*^, where *m* represents the dimensionality. Then *e*_1_, *e*_2_, …, *e*_|*C*|_ are the basic embeddings of the codes *d*_1_, *d*_2_, …, *d*_|*C*|_. Let *g*_*i*_ ∈ ℝ^*m*^ denote the final representation of the code *d*_*i*_, as a convex combination of the basic embeddings of itself and its ancestors:

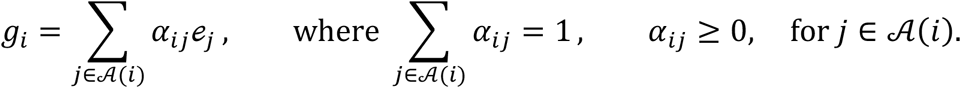

Here, 𝒜 (*i*) are the indices of the code *d*_*i*_ and 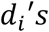 ancestors, *e*_*j*_ is the basic embedding of the code *d*_*j*_, and *α*_*ij*_ ∈ ℝ, the attention weight on the embedding *e*_*j*_ when calculating *g*_*i*_. The attention weight *α*_*ij*_ is calculated by the softmax function,

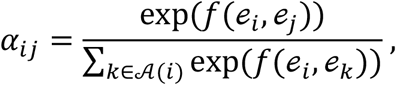

where *f*(*e*_*i*_, *e*_*j*_) is a scalar value representing the compatibility between the basic embeddings of *e*_*i*_ and *e*_*j*_, computed via the feed-forward network,

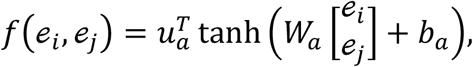

where *W*_*a*_ ∈ ℝ ^*I×*2*m*^ is the weight matrix for the concatenation of *e*_*i*_ and *e*_*j*_, *b*_*a*_ ∈ ℝ^*l*^ is the bias vector, and *u*_*a*_ ∈ ℝ^*l*^ is the weight vector for generating the scalar value, with *l* denoting the dimension size of the hidden layer *f*(*e*_*i*_, *e*_*j*_).

### End-to-end training with a predictive model

We use the above-described approach to train a model for predicting the disease risk at the next visit, denoted as *y*_*t* +1_ at time step *t* + 1 given all the previous visit history up to the current time step *ν*_1_, *ν*_2_, …, *ν*_*t*_,

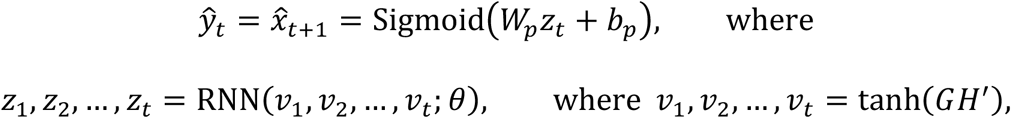

where we perform the sequential disease risk prediction using a recurrent neural network. An LSTM is used for the experiments in this work. The prediction loss is defined as

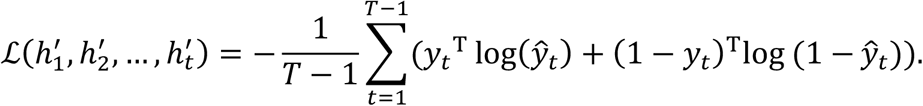

The pseudocode of the workflow of the ALIGATEHR training process is described in the Supplementary Information pseudo code.

### Experiments

We conduct experiments to assess if ALIGATEHR offers superior prediction performance compared to state-of-the-art baseline methods, ablation study, and to analyze the interpretability of ALIGATEHR.

Our aim is to predict whether a patient will receive a specific diagnosis during the next clinical visit. We compare ALIGATEHR with state-of-the-art models in the following categories:

1. Traditional machine learning methods: Logistic Regression (LR) and eXtreme Gradient Boosting (XGBoost). In both the LR and XGBoost models, the input feature vector, denoted as ***x***, is a list of the disease status gathered from all preceding visits for a given patient, i.e., the feature vector captures the historical disease information up to the current point in time.
2. Recurrent neural networks (RNN): Long Short-Term Memory. Input sequence *x*_1_, …, *x*_*t*_ of RNN consists of sequential record of disease status up to the current point in time for a patient, allowing the model to make predictions based on this longitudinal data.
3. The Skip-gram based Model: Med2Vec. Med2Vec follows the concept of Skip-gram, known for its simplicity and robustness in learning representations, attempting to construct meaningful representations of medical codes.
4. Attention-based models: GRAM and Dipole. In GRAM, the input sequence *x*_1_, …, *x*_*t*_ is first transformed by the embedding matrix, then fed to the Gated Recurrent Unit (GRU) with a single hidden layer, which in turn makes the prediction. In Dipole, an RNN-based risk prediction model that applies attention mechanism to perform visit analysis on top of bidirectional GRU, which can use attention weights to determine the importance of each visit.

To evaluate the robustness of our proposed model, we perform an ablation study to examine the influence of e-pedigrees and medical ontology graphs on the performance of ALIGATEHR. Our goal is to explore whether our model can effectively establish connections among crucial family members who significantly impact the patient’s disease risk. Specifically, we design the following setting:

1. ALIGATEHR^1^: Pedigree graph is removed. We only keep ontology graph in the model to assess the impact of family relations.
2. ALIGATEHR^2^: Medical ontology graph is removed. We only keep pedigree graph in the model to assess the impact of diagnoses from EHR.
3. ALIGATEHR^3^: Both pedigree graph and ontology graph are removed. A baseline model for comparison.
4. ALIGATEHR^4^: We maintain both pedigree graph and ontology graph, while specifically assign constant weights to the edges of the pedigree graph. The goal is to evaluate whether variations in contributions from distinct family members may influence the outcomes.
5. ALIGATEHR^5^: Similar to the above ablation setting, we specifically apply constant weights to the edges of the ontology graph only.
6. ALIGATEHR^6^: The model maintains constant weights on the edges of both pedigree graph and ontology graph.

For all models, we randomly partition the dataset into three parts, training data (70%), validation data (10%), and testing data (20%). In our experiments, the best-performing model on the validation data was selected and its performance was further evaluated on the test data in a 3-fold cross validation by calculating the area under a receiver operating characteristic curve (AUC).

## Data Availability

The data that support the findings of this study are available from IBM Watson Health, but restrictions apply to the availability of these data, which were used under license for the current study, and so are not publicly available.

## Data availability

The data that support the findings of this study are available from IBM Watson Health, but restrictions apply to the availability of these data, which were used under license for the current study, and so are not publicly available. Data are however available from the authors upon reasonable request and with permission of IBM Watson Health.

## Code availability

The source code of ALIGATEHR is publicly available at: https://github.com/XiayuanHuang/ALIGATEHR

## Acknowledgements

This work was supported by the Yale-BI biomedical data science fellowship.

## Author contributions

X.H., Z.D., Z.W. and J.d.J. conceived the study. X.H. implemented the algorithm, conducted the experiments, and performed all analyses. Z.W. and J.d.J. supervised the study. J.A., A.M.E, D.L. and H.Z. provided input on analysis, presentation and/or model interpretability. X.H., Z.W. and J.d.J. wrote the manuscript. All authors provided feedback and approved the manuscript.

## Additional information

### Extended data

**Extended Data Fig. 1.**
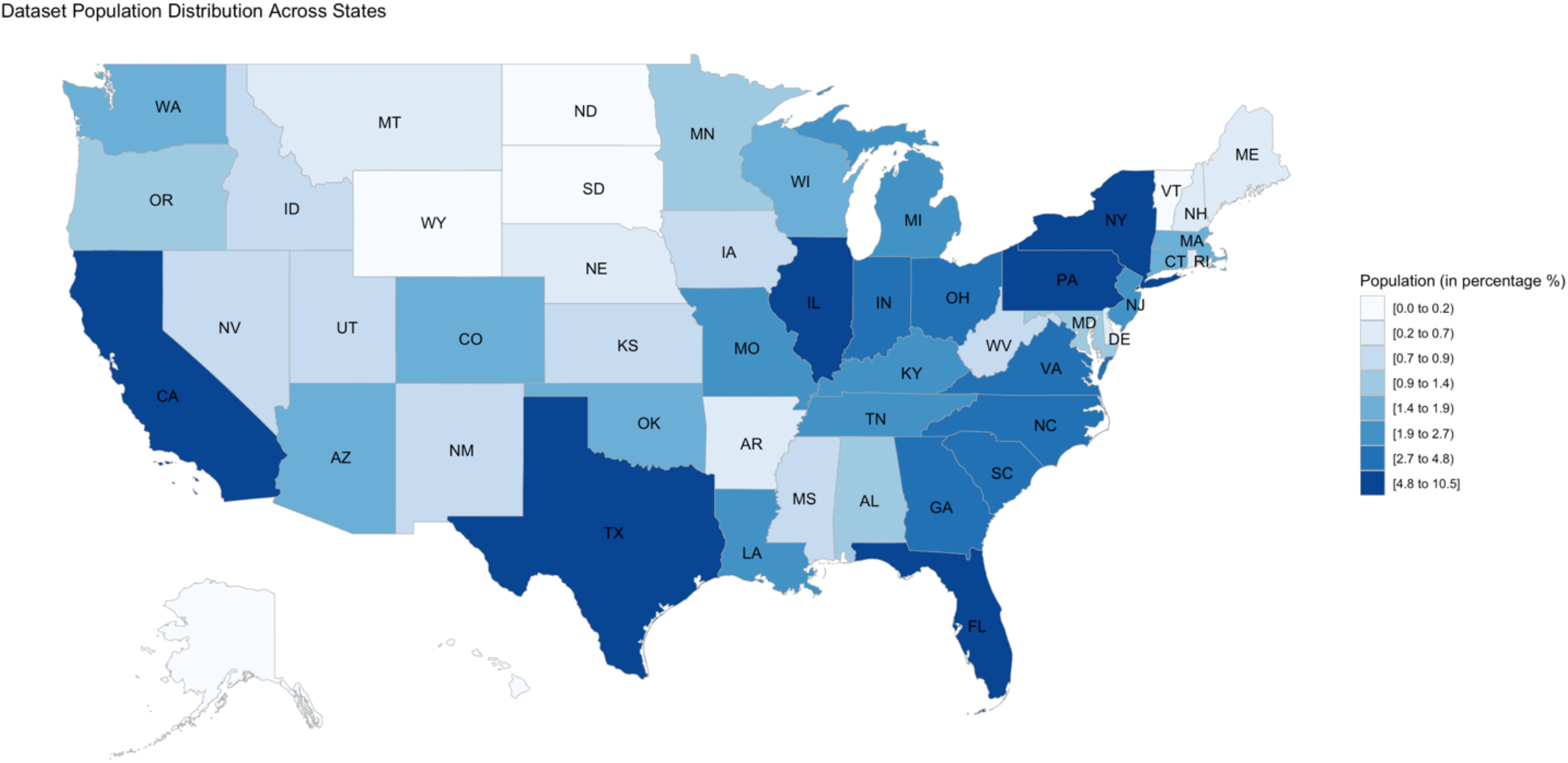
Distribution of population percentages across states in the experimental EHR dataset.

**Extended Data Table 1.**
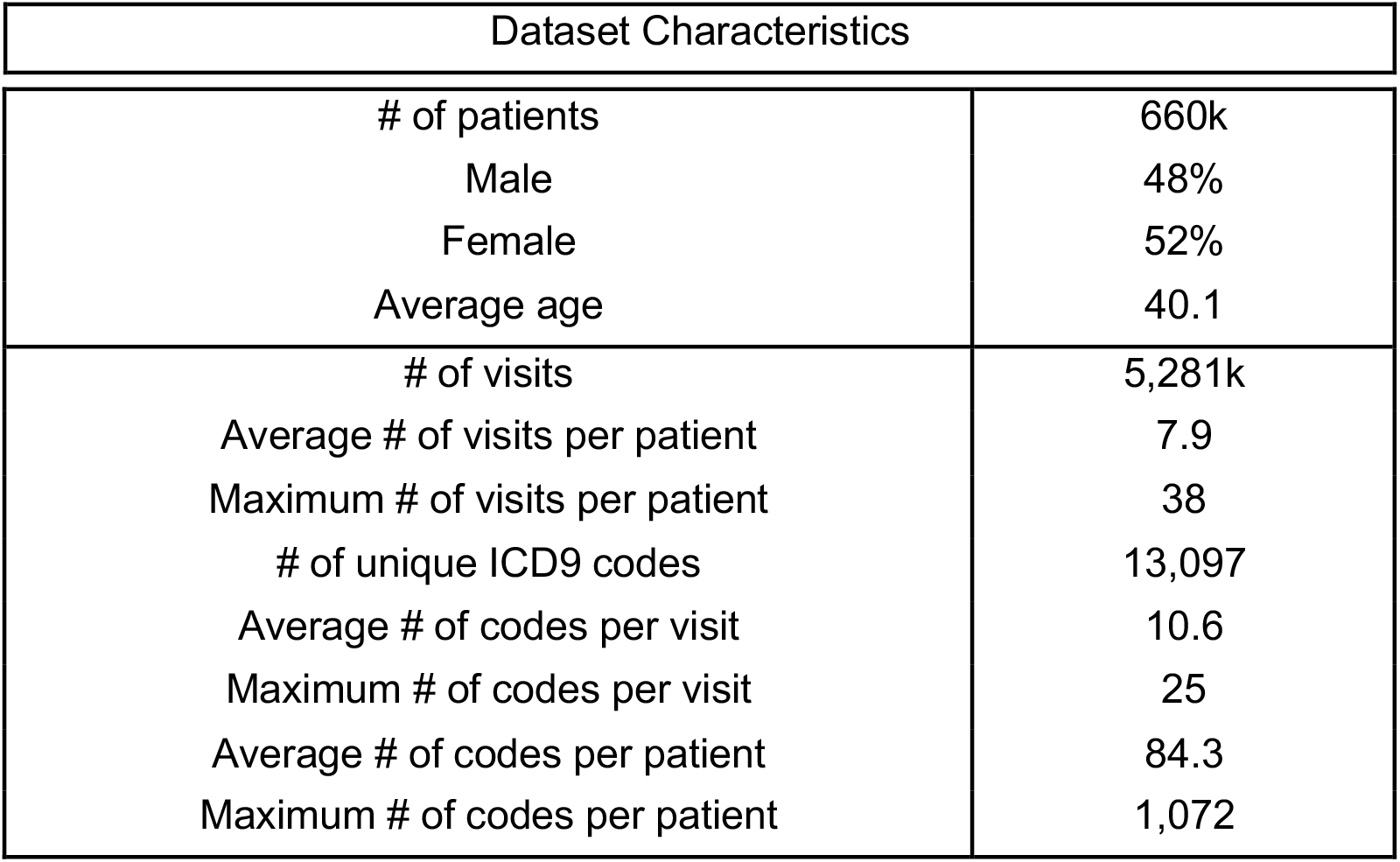
Basic characteristics of the experimental EHR dataset

| Dataset Characteristics |  |
| --- | --- |
| # of patients | 660k |
| Male | 48% |
| Female | 52% |
| Average age | 40.1 |
| # of visits | 5,281k |
| Average # of visits per patient | 7.9 |
| Maximum # of visits per patient | 38 |
| # of unique ICD9 codes | 13,097 |
| Average # of codes per visit | 10.6 |
| Maximum # of codes per visit | 25 |
| Average # of codes per patient | 84.3 |
| Maximum # of codes per patient | 1,072 |

**Extended Data Table 2.** Notations in ALIGATEHR

| Layer | Notation | Description |
| --- | --- | --- |
| Knowledge graph | $D$ | Set of leaf codes |
| | $D'$ | Set of internal nodes |
| | $C$ | Set of entire diagnose codes |
| | $V_t$ | A patient's visit at time step $t$ |
| | $X_t \in \{0,1\}^{\vert C\vert }$ | A vector representation of $V_t$ |
| E-pedigrees graph | $h_i$ | Input vector of patient $i$'s disease representation |
| | $h'_i$ | Vector of patient $i$'s disease latent representation |
| | $\sigma(\cdot)$ | An activation function that applies nonlinearity |
| | $\alpha_{ij}$ | Attention score |
| | $W_e \in \mathbb{R}^{\vert C \times C\vert }$ | A weighted matrix for linear transformation |
| Predictive neural networks | $\hat{y}_t \in \mathbb{R}$ | Disease label output |
| | $z_t$ | RNN's hidden layer at the $t$-th time step |
| | $v_t \in \mathbb{R}^{\vert C\vert }$ | $t$-th visit representation |
| | $W_p \in \mathbb{R}^{\vert C\vert }$ | Weight matrix of Softmax function |
| | $b_p$ | Bias of Softmax function |
| | $\theta$ | Parameters of RNN |
| | $\mathcal{L}(\cdot)$ | Entropy loss function |

## Supplementary information

**“Boehringer Ingelheim – Global Computational Biology and Digital Sciences” authors:** Author list: Jatin Arora, Abdullah Mesut Erzurumluoglu, Daniel Lam, Pierre Khoueiry, Jan N Jensen, James Cai, Nathan Lawless, Jan Kriegl, Zhihao Ding, Johann de Jong

Address: Global Computational Biology and Digital Sciences (gCBDS), Boehringer Ingelheim Pharma GmbH & Co. KG, Biberach an der Riss, Germany

**Pseudo code:** ALIGATEHR algorithm

### Algorithm 1

ALIGATEHR Pseudocode

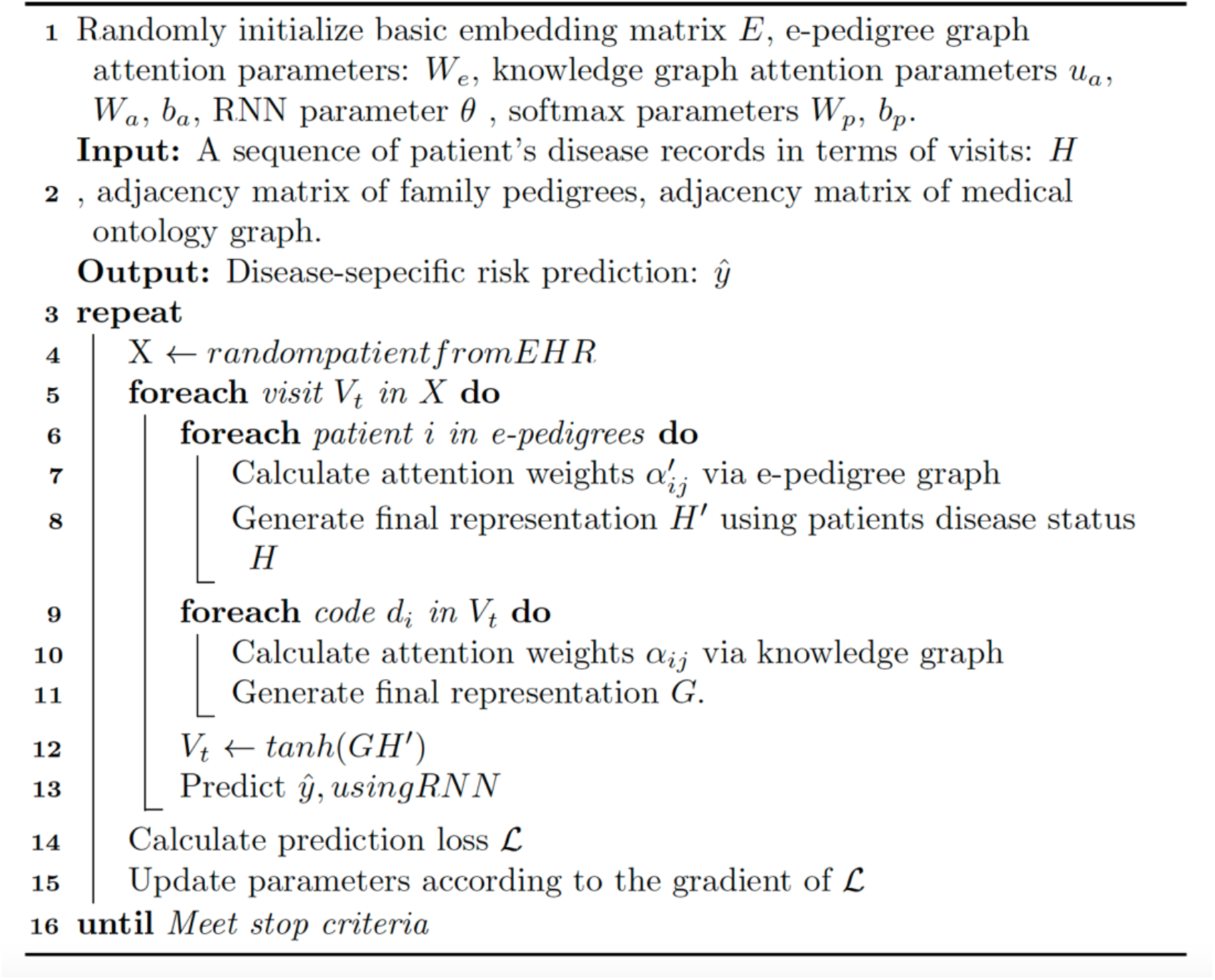

